# Does the Pareto Principle describe parasite counts in humans? The case of hookworm in pregnant women

**DOI:** 10.1101/2020.12.30.20249036

**Authors:** Yenifer Orobio, Neal Alexander

## Abstract

The Pareto Principle (PP) — that 80% of an attribute are found in 20% of individuals — is one way to characterize heterogeneity in infectious diseases. An alternative is the dispersion parameter (*k*) of the negative binomial distribution (NBD). The NBD has two parameters, while the PP is a single property which may hold for any distribution family. The objectives of the current work are: a) to obtain a relation between the PP and NBD, i.e. to specify which combinations of NBD parameters comply with the PP; b) for hookworm, a soil-transmitted intestinal parasite of humans, to identify whether the PP or the NBD is a more parsimonious description of heterogeneity of infection load. For objective a), an empirical relation is found in the form of a saw-toothed line on a plot of *k* against the mean, reaching an asymptote of approximately 0.24. For objective b), we estimate *k* and the mean from nine studies from a systematic review of hookworm in pregnant women. Seven studies had higher heterogeneity than the PP, ranging from 83:20 to 100:20: we call these super-Pareto. One study was sub-Pareto (74:20), and one was Pareto (80:20). This suggests that at least two parameters, as supplied by the NBD, are necessary to describe the heterogeneity of hookworm. The probability of reaching a target reduction in prevalence is less when there is greater aggregation, which suggests that estimating aggregation via a subsample could be worthwhile, in order to set a target coverage threshold before starting mass drug administration.

## Introduction

Infectious disease processes are typically heterogeneous, as reflected in phenomena such as ‘superspreaders’, i.e. a small proportion of hosts being responsible for a large proportion of transmission (Jankowski et al. 2013; Lloyd-Smith et al. 2005; Stein 2011). One way to characterize such heterogeneity is the Pareto principle, also known as the 80/20 rule, which states that 80% of a certain characteristic is harbored by 20% of the population (Anderson and May 1991; Hardy 2010; Holland 2013). This has been used to describe the distribution of transmission and spread of infection among hosts of a variety of diseases (Cooper et al. 2019; Stein 2011; Woolhouse et al. 1997). In the case of count data, another way to characterize heterogeneity is via the dispersion parameter (*k*) of the negative binomial distribution (NBD), which is a generalization of the Poisson (Lloyd-Smith 2007). The NBD has often been found to well describe host-level patterns of helminths and other parasites (Anderson and May 1991). The NBD has two parameters (*k* and the mean, *μ*), while the PP is a single numerical property which may hold for any distribution family.

In the current paper we compare the ability of the PP and the NBD to characterize heterogeneity in infection intensity of hookworm, specifically hookworm in pregnancy. Hookworm is a soil-transmitted intestinal helminthiasis, which the World Health Organization (WHO) considers a major cause of morbidity, closely linked to poverty and inadequate personal hygiene, consumption of raw foods, fecal contamination of the environment and lack of sanitary services and drinking water (Martínez de la Ossa et al. 2010; Pawlowski et al. 1992; Valerio et al. 2002). The human hookworm pathogens are *Necator americanus* and *Ancylostoma duodenale*. The resulting morbidity is associated with infection intensity (Organización Panamericana de la Salud 2012), so epidemiological descriptions need to reflect the distribution of numbers of parasite between hosts, as well as prevalence. Intense infections cause chronic blood loss that can lead to physical, nutritional and cognitive deterioration, specifically growth retardation and anemia in children and pregnant women (Organización Mundial de la Salud 2005; Organización Panamericana de la Salud 2012; Roca et al. 2003).

The objectives of the current work are: a) to obtain a relation between the PP and NBD, i.e. to specify which combinations of NBD parameters comply with the PP; b) to determine, in the case of hookworm in pregnancy, whether the PP or the NBD is a more parsimonious description of heterogeneity.

## Materials and Methods

### Comparison of the negative binomial distribution and the Pareto principle

Some combinations of NBD parameters give a lower degree of heterogeneity, e.g. 70:20 rather than 80:20, and we call this *sub-Pareto*. Other combinations give a higher degree, e.g. 90:20, which we call *super-Pareto*, while we will call a distribution in accordance with the Pareto principle *equi-Pareto*. The strategy of the current paper is to fit the negative binomial to observed data, read off whether this fitted distribution is *sub*-, *super*- or *equi-Pareto*, and so determine whether or not the Pareto is a parsimonious description of the heterogeneity of the data.

### Negative binomial distributions which satisfy the Pareto Principle

We use the following procedure to identify those combinations of the negative binomial dispersion (*k*) and mean (*μ*) parameters which meet the PP. Values of *μ* were considered between 1 and 5000: more specifically, a geometric series of 1000 values between these limits, i.e. the log-values were uniform between log(1) and log(5000). Then, for each value of *μ*, the value of *k* was identified which satisfied the PP. To do so, the ‘nlminb’ function of R optimized over the proportion of the total count found in the highest 20 percent of the values. The cumulative distribution of the negative binomial was obtained as the cumulative sum of its probability function (R Core Team 2019; Venables and Ripley 2002). Hence we define an empirical, non-algebraic, relation between those values of *μ* and *k* which satisfy the PP constraint.

### Case study: hookworm in pregnancy

Hookworm in humans is caused by two species of nematode parasites: *Necator americanus* and *Ancylostoma duodenale*, whose larvae infect people via contact with soil (Hotez et al. 2004). Disease results largely from blood loss caused by the adult parasites in the intestine. The morbidity largely results from anemia, and is associated with the intensity of infection. Almost 500 million people in developing tropical countries are infected (Loukas et al. 2016), including a quarter to a third of pregnant women in sub-Saharan Africa. The latter estimates are from a systematic review of hookworm-related anemia among pregnant women (Brooker et al. 2008), whose data we use in the current paper. The variables extracted from the systematic review were: sample size of the study population, proportion of people with infection, initial prevalence of infection, infection intensity category.

The original studies report hookworm eggs per gram (epg), which are obtained by applying a conversion factor to the numbers of eggs counted under the microscope. For example, one Kato Katz slide usually contains approximately 1/24 g of feces, so, if basing the eggs per gram on a single slide, the egg count would be multiplied by 24 (Organización Mundial de la Salud 1994; World Health Organization 1992). To apply the NB distribution, we need to infer the original egg counts which take values 0, 1, 2, etc., as opposed to the eggs per gram values which would be 0, 24, 48 etc. This factor will vary according to the technique (e.g. Kato-Katz or McMaster), the numbers of slides read per sample, and the value assumed for the mass of feces per slide. This factor was taken or inferred from each article, as detailed in **S1 Table**.

### Estimation of negative binomial parameters from grouped data

The data are available as the number of subjects within each category of infection intensity. For example, the World Health Organization defines light, moderate and heavy intensity infections as 1-1,999, 2,000-3,999 and ≥ 4,000 epg, respectively (Montresor et al. 2002; World Health Organization 2012). However, some publications used different categories. To estimate the two parameters of the negative binomial distribution from such data, the following procedure was used. Within each category, the probability of each possible value in that range occurring was calculated, as a function of the two parameters (*μ* and *k*), and summed to give a probability per category. There is no upper bound for the most intense category, so, to avoid an infinite sum, this probability was calculated as 1 minus the sum of the probabilities of the lower categories. The overall log-likelihood was then calculated by multiplying, within each category, the number of observations by the logarithm of the probability, then summing this product over categories. Finally, the likelihood was maximized over values of *μ* and *k* to obtain the fitted values of these parameters. This method requires at least three categories of infection intensity with positive numbers of women. This method was checked by generating simulated data, categorizing them, and ensuring that the original parameter values were recovered.

## Results and discussion

### Comparison of the negative binomial distribution and the Pareto principle

The pairs of negative binomial mean (*μ*) and dispersion (*k*) parameters which comply with the Pareto principle are shown in Fig 1. As the mean increases, the corresponding value of *k* follows a sawtooth, but generally declining, pattern, towards an asymptote of 0.244. The complexity of this sawtooth pattern is presumably related to the discrete nature of the distribution, and seems to preclude it being expressed exactly by a simple equation.

**Fig 1.**
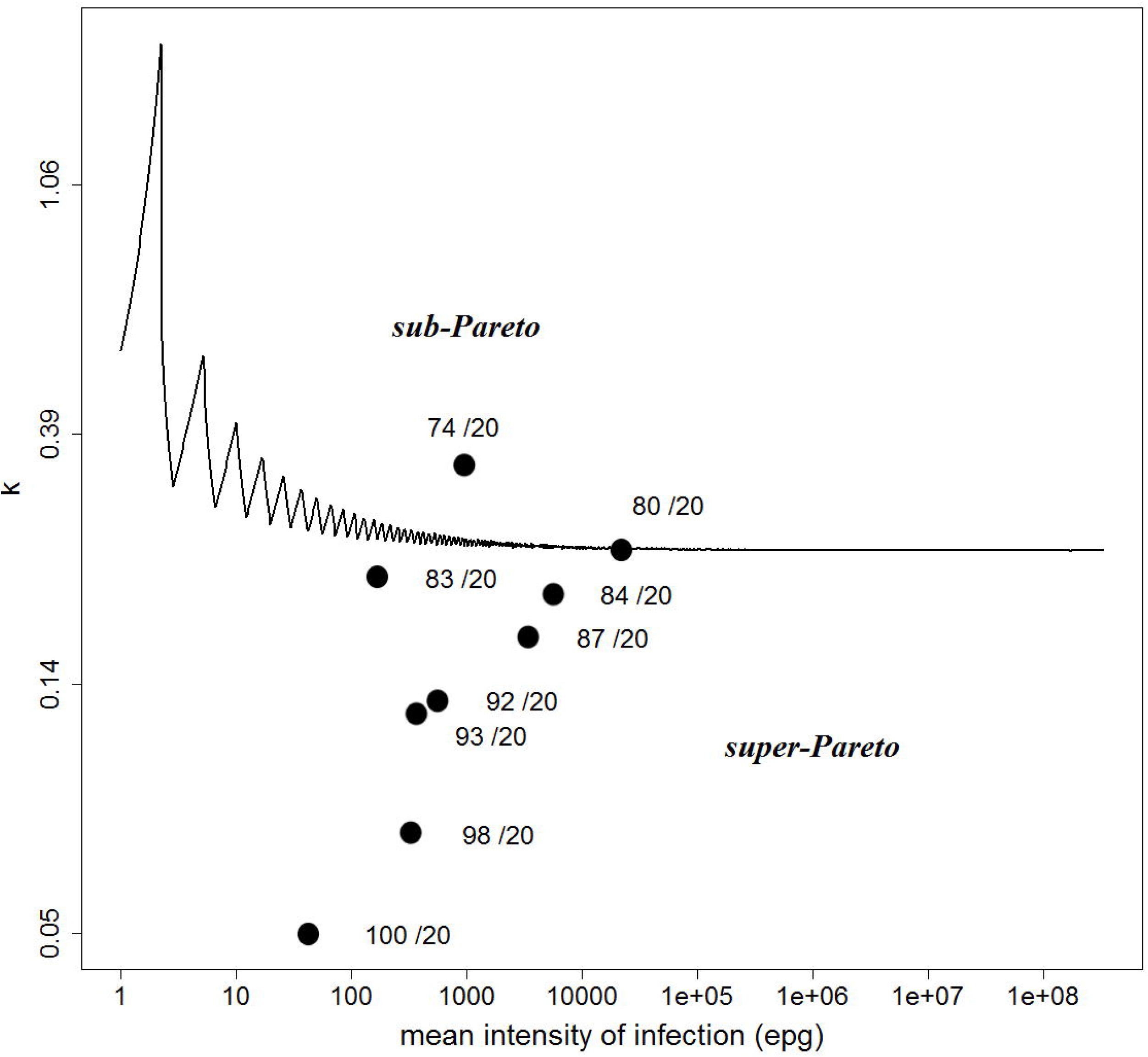
Relation between the Pareto principle, parameters of the negative binomial distribution, and aggregation of hookworm. The black line represents those combinations of the parameters of the negative binomial distribution which agree with the Pareto principle. This means that, on the black line, 80% of parasites (or any other attribute) will be found in 20% of people. The vertical axis for *k*, and the horizontal for the mean (*μ*) are both logarithmic. Each orange dot represents the negative binomial parameters from one study of hookworm in pregnancy: the majority are below the black line, indicating a greater degree of clustering than indicated by Pareto principle.

The Pareto principle (PP) affirms that the Lorenz curve (Butler and McDonald 1989) passes through the 80:20 point. For values of the mean in the range of *R*_0_ for various infections, values of *k* around 0.3 satisfy the PP (Lloyd-Smith et al. 2005; Susswein and Bansal 2020). Here we have found such *k* values over a wider range of values of the mean, while taking into account the discrete nature of the NBD. We also sought an explicit expression for the curve in Fig 1, and hence for the value of its asymptote, using generalized gamma, and lognormal, approximations to the negative binomial distribution. For the former, Butler and McDonald (1989) provide an approximation to the normalized incomplete moment and state, moreover, that this itself has another generalized gamma distribution. However, in general, the first moment does not have a closed form solution (Evans et al. 2000). Turning to the lognormal, such an approximation is appealing because, for large *μ*, the variance of the negative binomial is close to *μ*^2^/*k*. This means that, for constant *k*, the negative binomial distribution has constant coefficient of variation (i.e. ratio of the standard deviation to the mean), a property shared with the lognormal distribution. Moreover, the Lorenz curve for the lognormal has a very simple form (Krause 2014). However, equating the first two moments of the two distributions does not give a good approximation, nor does the suggestion of Best and Gipps (Best and Gipps 1974) to equate the second and third moments (not shown). Hence we have not been able to find an explicit expression for the asymptote in Fig 1.

### Case study: hookworm in pregnancy

**S1 Table** shows that the mean (*μ*) and dispersion (*k*) parameters could be estimated in 9 of the 13 (70%) component studies in the review, these being the ones with three or more categories of infection intensity. **S1 Fig** shows that these fitted distributions give a qualitatively good fit to the observed data. However, a limitation of this analysis is the inclusion of one study (Sill et al. 1987) which used a modified Kato technique with Ovo-FEC kits, which is semi-quantitative. The 9 pairs of parameter estimates are shown as points in Fig 1. Seven studies showed super-Pareto variation, i.e. more than 80% of the parasites in 20% of the population (below the line in Fig 1), one was *sub-Pareto* (less than 80% of the parasites in 20% of the population; above the line in Fig 1), and one was *equi-Pareto*. Hence, the parasite distribution in these studies tended to be even more concentrated than implied by the Pareto principle.

The Pareto principle has been proposed as a guide to identifying priority control areas for parasitic diseases (Pan American Health Organization 2019). Although we have found that hookworm infections in pregnant women, at the individual level, are usually super-Pareto, other parasites and insects may be nearly equi-Pareto. For example, this has been reported for ectoparasites, and specifically ticks (Warwick et al. 2016). For disease vectors, a recent study found that bloodfed *Anopheles* mosquitoes per individual were equi-or super-Pareto (Guelbeogo et al. 2018). For phlebotomine vectors of leishmaniasis, a recent study (Moreno et al. 2020) analyzed abundance at five sites in Colombia, using negative binomial models, and the numbers per house were super-Pareto only two of the sites and the other three sites were sub-Pareto. Overall, it is important for control programs to reach those with highest parasite densities or vector exposure, including potential superspreaders (Cooper et al. 2019; Harrison and Bennett 2012).

## Conclusions

The case study of hookworm infection in pregnant women did not support the use of the Pareto principle as an empirical description of parasites clustering within people. Rather, in most component studies of the systematic review, the clustering was super-Pareto, i.e. with greater aggregation than stated by the principle.

The negative binomial distribution was the alternative model, and, for this, we developed a method for estimating its parameters from grouped data, in particular the data tables provided in the component studies of the systematic review. We also found, empirically, the relation between those values of the negative binomial distribution’s mean and dispersion parameters for which the Pareto principle holds. We were not able to find a closed form expression for the line that embodies these pairs of values, although future work could exploit the relation between the Pareto principle and the Lorenz curve of ecology.

Finally, the existence of super-Pareto aggregation emphasizes the need for control programs of parasitic diseases to reach those few people who harbor a high proportion of the parasites.

## Data Availability

All data are included in the manuscript and its supporting information.

## Acknowledgments

We appreciate Simon Brooker providing information from the systematic review work on pregnant women. Karim Anaya-Izquierdo helped orient us in our pursuit of an explicit expression for the Lorenz curve. This article includes findings from a master’s degree in Epidemiology conducted by Yenifer Orobio at the School of Public Health of the Universidad del Valle in Cali, Colombia, and we thank the tutors and assessors for their guidance during this degree project. We are also grateful to multiple statistician and epidemiologist colleague, whose ingenious ideas which helped structure the research question and approach, and to the evaluators of the degree project, whose questions helped enrich the discussion.

## Declarations

### Availability of data and material

All data are included in the manuscript and its supporting information.

### Ethics approval

This study was reviewed by the Human Ethics Committee of the Faculty of Health of Universidad del Valle (approval number 083-015).

