## Supplemental Figure 1 for "Does the Pareto Principle describe parasite counts in humans? The case of hookworm in pregnant women"

**S1 Fig. Histogram of egg count per gram of hooks grouped in each study of the systematic review.**

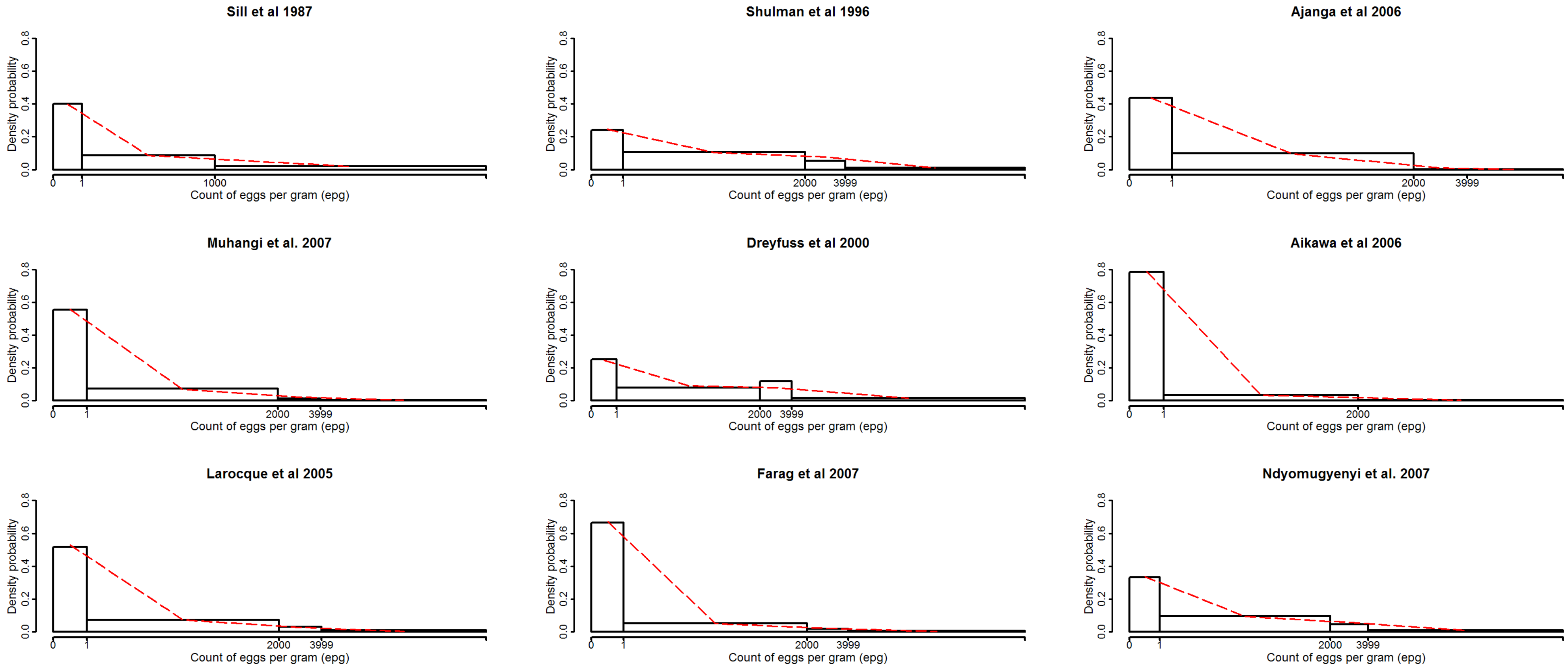


The axis labels show the original values, although the scale was transformed with the fourth root. The cropped line (red) represents the fitted negative binomial distribution
