## Supplemental Table 1 for "Does the Pareto Principle describe parasite counts in humans? The case of hookworm in pregnant women"

**S1 Table. Estimation of dispersion parameters *k* and mean intensity of infection *μ* from grouped data of the intensity of infection in each study of the systematic review by Brooker et al (19).**

| Authors (Study) | Study area | Target population | Date | Sample size | Proportion of people with infection (%) | Classes of intensity of infection for hookworms (epg [a]) (22) | | | | Number of subjects infected by categories of intensity for hookworms | | | | Factor of multiplication for egg counts estimation | Technique for egg counts estimated per gram | Initial value of the mean of total eggs seen | Mean intensity of infection (epg*) ($\mu$) | Dispersion parameter ($k$) | Deviance (D) |
|---|---|---|---|---|---|---|---|---|---|---|---|---|---|---|---|---|---|---|---|
| | | | | | | No infection | Light | Moderate | Heavy | No infection | Light | Moderate | Heavy | | | | | | |
| Glover-Amengor, 2005 (36) | Ashanti Region, Ghana | Women attending antenatal clinic aged 15–49, 2003–2005 | 2004 | 86 | 8.1 | 0 | 1-1,999 | | | 79 | 7 | | | - | not reported | | - [d] | | |
| Jackson, 1987 (37) | Monrovia, Liberia | Women attending antenatal clinic aged 14–43, 1985 | 1986 | 128 | 38.0 | 0 | 1-1,999 | | | 90 | 38 | | | - | not reported | | - [d] | | |
| Rodriguez-Morales, 2006 (38) | Nine states, Venezuela | Women attending antenatal clinic aged 18–42, 2003–2004 | 2004 | 1038 | 8.1 | 0 | 1-1,999 | | | 271 | 767 | | | - | Kato-Katz | | - [d] | | |
| Ayoya, 2006 (39) | Bamako, Mali | Women attending antenatal clinic | 2002 | 131 | 8.2 | 0 | 1-1,999 | | | 121 | 10 | | | - | Kato-Katz | | - [d] | | |
| Aikawa, 2006 (40) | Yen Thanh district, Vietnam | Community-based sample | 2003 | 391 | 21.4 | 0 | 1-1,999 | 2000-3999 | ≥ 4,000 | 307 | 78 | 6 | | 24 [b] | Kato-Katz | 2.76 | 5.08 | 0.0528 | 450.10 |
| Farag, 2007 (41) | Pemba Island, Zanzibar (Tanzania) | Women attending antenatal clinic | 2004 | 970 | 32.9 | 0 | 1-1,999 | 2000-3999 | ≥ 4,000 | 651 | 278 | 24 | 17 | 24 [b] | Kato-Katz | 10.27 | 12.38 | 0.0787 | 1529.18 |
| Muhangi, 2007 (42) | Entebbe, Uganda | Women attending antenatal clinic | 2005 | 2498 | 44.5 | 0 | 1-1,999 | 2000-3999 | ≥ 4,000 | 1386 | 1025 | 44 | 43 | 24 [b] | Kato-Katz | 9.10 | 12.79 | 0.1268 | 4183.02 |
| Larocque, 2005 (43) | Iquitos, Peru | Women attending antenatal clinic aged 18–42, 2003–2004 | 2003 | 1042 | 47.2 | 0 | 1-1,999 | 2000-3999 | ≥ 4,000 | 550 | 435 | 40 | 17 | 24 [b] | Kato-Katz | 12.28 | 15.54 | 0.1344 | 1863.89 |
| Sill, 1987 (28) | Papua New Guinea | Women attending antenatal clinic, 1985 | 1985 | 30 | 60.0 | 0 | 1-999 | 1000+ | | 12 | 12 | 6 | | 24 [b] | Kato-Katz | 17.20 | 34.29 | 0.1731 | 63.30 |
| Ndyomugyenyi, 2008 (44) | Masindi, Uganda | Women attending antenatal clinic aged 14–42, 2003–2004 | 2004 | 832 | 66.6 | 0 | 1-1,999 | 2000-3999 | ≥ 4,000 | 274 | 456 | 46 | 52 | 20 [c] | Kato-Katz | 30.80 | 42.40 | 0.2055 | 1708.06 |
| Ajanga, 2006 (45) | Ukerewe Island, Tanzania | Women attending antenatal clinic aged 15–45, 2004 | 2004 | 972 | 56.4 | 0 | 1-1,999 | 2000-3999 | ≥ 4,000 | 424 | 538 | 4 | 6 | 24 [b] | Kato-Katz | 3.30 | 9.22 | 0.2195 | 1462.74 |
| Dreyfuss, 2000 (46) | Sarlahi district, Nepal | Community-based sample of micronutrient supplement in women aged 15-40, 1994-1997 | 1999 | 336 | 74.2 | 0 | 1-1,999 | 2000-3999 | ≥ 4,000 | 49 | 88 | 29 | 24 | 24 [b] | Kato-Katz | 68.14 | 75.77 | 0.2446 | 482.62 |

| | | | | | | | | | | | | | | | | | | |
|---|---|---|---|---|---|---|---|---|---|---|---|---|---|---|---|---|---|---|
| Shulman, 1996 (47) | Kilifi, Kenya | Women attending antenatal clinic with aged 15-41, 1993 | 1993 | 251 | 74.9 | 0 | 1-1,999 | 2000-3999 | ≥ 4,000 | 63 | 162 | 18 | 18 | 50 [b] | McMaster | 35.17 | 19.51 | 0.3439 | 529.65 |

[a] epg: eggs per gram of faecal material

[b] Two slides of 41.7 mg of faecal material per sample were assumed (22). In case of the McMaster method (48) being used, rather than Kato-Katz, a multiplication factor of 50 (rather than 24) was inferred for the total number of eggs.

[c] Reported in the article.

[d] Not estimated because at least three categories of infection intensity are required.
